# Three-Dimensional Study on Pharyngeal Morphology in Mouth-Breathing Children

**DOI:** 10.1101/2025.08.24.25334325

**Authors:** Siting Zhu, Xiao He, Bin Hu, Jiying Li

**Author notes:** Correspondence to: Jiying Li Dancun Road, 530031, Nanning ChinPhone.

## Abstract

**Objectives:** To investigate the pharyngeal morphological differences between mouth-breathing and nose-breathing children.

**Material and Methods:** Sixty-two children aged 8–10 years underwent cone-beam computed tomography (CBCT) examinations. They were divided into two groups based on breathing mode: mouth-breathing group (30 cases; males: 22, females: 8) and nose-breathing group (32 cases; males: 20, females: 12). The pharyngeal cavities were reconstructed three-dimensionally using Mimics software, with measurements of pharyngeal segment volume and length. Independent-samples t-tests were used for parameter comparisons.

**Results:** Total and segmental pharyngeal volumes were significantly smaller in the mouth-breathing group than in the nose-breathing group (P < 0.05), whereas pharyngeal length showed no significant difference (P > 0.05). Additionally, the volume-to-length ratio in the mouth-breathing group was significantly lower than in the nose-breathing group (P < 0.05).

**Conclusions:** Mouth-breathing children exhibit a significantly narrower pharyngeal morphology, characterized by reduced volume without changes in length, leading to decreased airway patency. This structural alteration correlates with increased respiratory resistance and has often been linked to maxillary constriction and posterior pharyngeal space reduction.

## INTRODUCTION

Breathing is a fundamental physiological function essential for maintaining life^[1]^. Abnormal breathing patterns represent a significant public health issue affecting children’s healthy development. Mouth-breathing (MB) is defined as breathing through the mouth for 5–30% of the time during a period exceeding six months^[2]^. Epidemiological studies indicate that approximately 12–55% of school-aged children exhibit persistent mouth-breathing habits^[3–6]^, with 60–70% associated with upper airway obstructions such as adenoid hypertrophy, allergic rhinitis, and nasal septum deviation^[2, 7, 8]^. Chronic mouth breathing can lead to abnormal tongue posture, mandibular retraction, and increased vertical craniofacial development — manifesting as characteristic "mouth-breathing facial morphology." The core pathological mechanism underlying these changes is closely linked to morphological alterations in pharyngeal anatomy^[9–12]^.

The pharyngeal cavity, as a critical anatomical region of the respiratory tract, directly influences respiratory dynamics. Anatomically divided into nasopharynx, velopharyngeal, and glossopharyngeal regions, these subdivisions are demarcated by soft-tissue structures like the adenoids, palatine tonsils, and tongue^[13]^. Previous 2D cephalometric studies revealed increased facial height and elevated hyoid position in mouth-breathing children^[14]^. Nevertheless, 2D imaging fails to comprehensively capture complex three-dimensional structural changes^[12, 15]^. Advancements in CBCT and 3D reconstruction technology have made precise quantification of pharyngeal morphology achievable. Mimics software enables volumetric segmentation and surface reconstruction to visualize 3D anatomy, allowing accurate measurements of linear distances, cross-sectional areas, and volumes.

Current research on pharyngeal morphology in mouth-breathing children primarily focuses on single anatomical subregions or cross-sectional analyses^[16, 17]^. Comparative 3D morphological studies across contiguous naso-velo-glosso-pharyngeal regions remain limited^[12]^. Clarifying morphological characteristics of pharyngeal subregions in mouth-breathing children and their divergence from nasal breathers would not only provide anatomical evidence for the "mouth-breathing-craniofacial dysfunction" pathological axis but also offer precise morphological guidance for early interventions like adenoidectomy, orofacial myofunctional therapy, or orthodontic treatment^[18, 19]^. Therefore, this study employs CBCT and Mimics-based 3D reconstruction to compare morphological parameters of nasopharyngeal, velopharyngeal, and glossopharyngeal regions between mouth-breathing and nose-breathing children, aiming to elucidate the impact of oral respiration on pediatric pharyngeal structure and inform clinical precision medicine.

## MATERIAL AND METHODS

### Research subjects

A total of 62 children who visited the Department of Stomatology at the Third Affiliated Hospital of Guangxi Medical University from January 2023 to December 2023, were selected, including 42 males and 20 females. They were divided into two groups based on their breathing patterns: the mouth-breathing group consisting of 30 participants (22 boys and 8 girls) and the nose-breathing group consisting of 32 participants (20 boys and 12 girls).

Inclusion criteria were as follows:

1. Age 8–10 years old;
2. Body Mass Index (BMI) within the normal range (13.8–19.1 for boys and 13.5–19.0 for girls);
3. No bad habits such as thumb-sucking, lip-biting, or tongue protrusion;
4. No systemic diseases of the heart, lungs, or nervous system.

Exclusion criteria were as follows:

1. Previous or current orthodontic treatment;
2. Nasal diseases or history of nasal surgery;
3. Facial deformities;
4. Diagnosis of Obstructive Sleep Apnea Syndrome (OSAS);
5. Upper respiratory tract infection during CBCT scanning;
6. Incomplete or inadequate case and examination data.

This study was approved by the Medical Ethics Committee of the Third Affiliated Hospital of Guangxi Medical University (Nanning Second People’s Hospital), Ethics Approval Number: Y2022018. All guardians of the study participants were informed about the study and provided informed consent to participate.

## Methods

### Breathing mode classification

This study employed a combination of subjective assessment (questionnaire survey and clinical interview) and objective detection (parent self-assessment and clinical re-examination) to group the research subjects, aiming to minimize misclassification bias. The specific procedures are as follows. At the first visit, patients with suspected mouth breathing were initially screened through clinical interviews and completing questionnaires, which included basic information including the patient’s name, gender, date of birth, height, and weight, as well as a mouth breathing symptom screening scale (Table 1). Then, parents were taught to use the mirror check or cotton wool methods to examine their children for mouth breathing, and videos were periodically recorded for clinical assessment by doctors. At least six months later, a re-examination was conducted for clinical verification. Within three minutes, a double-sided mirror was placed under the nostrils while the patient sat in a resting position. The presence of fog on both sides of the mirror was observed to assess the breathing type of each patient.

**Table 1.**
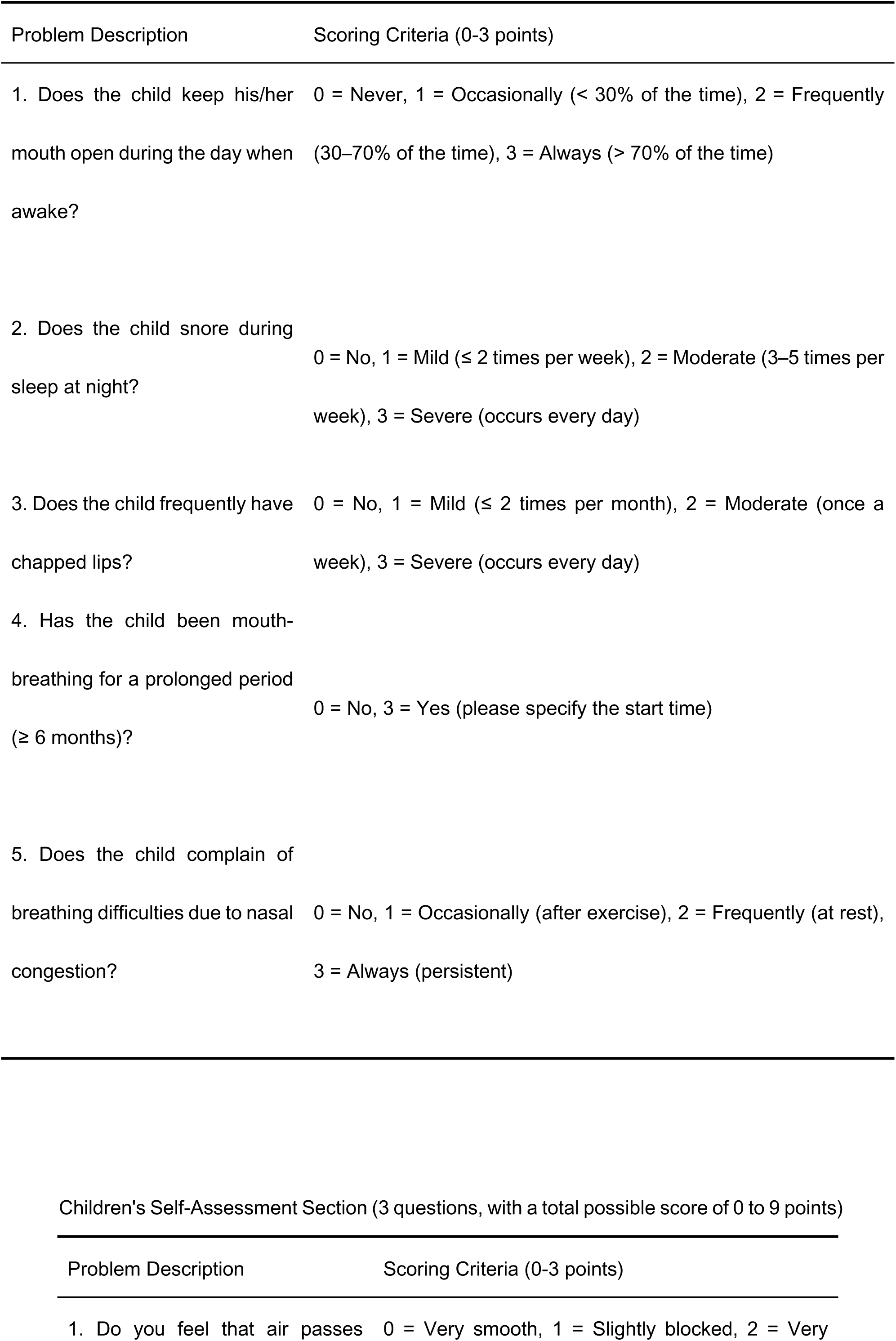
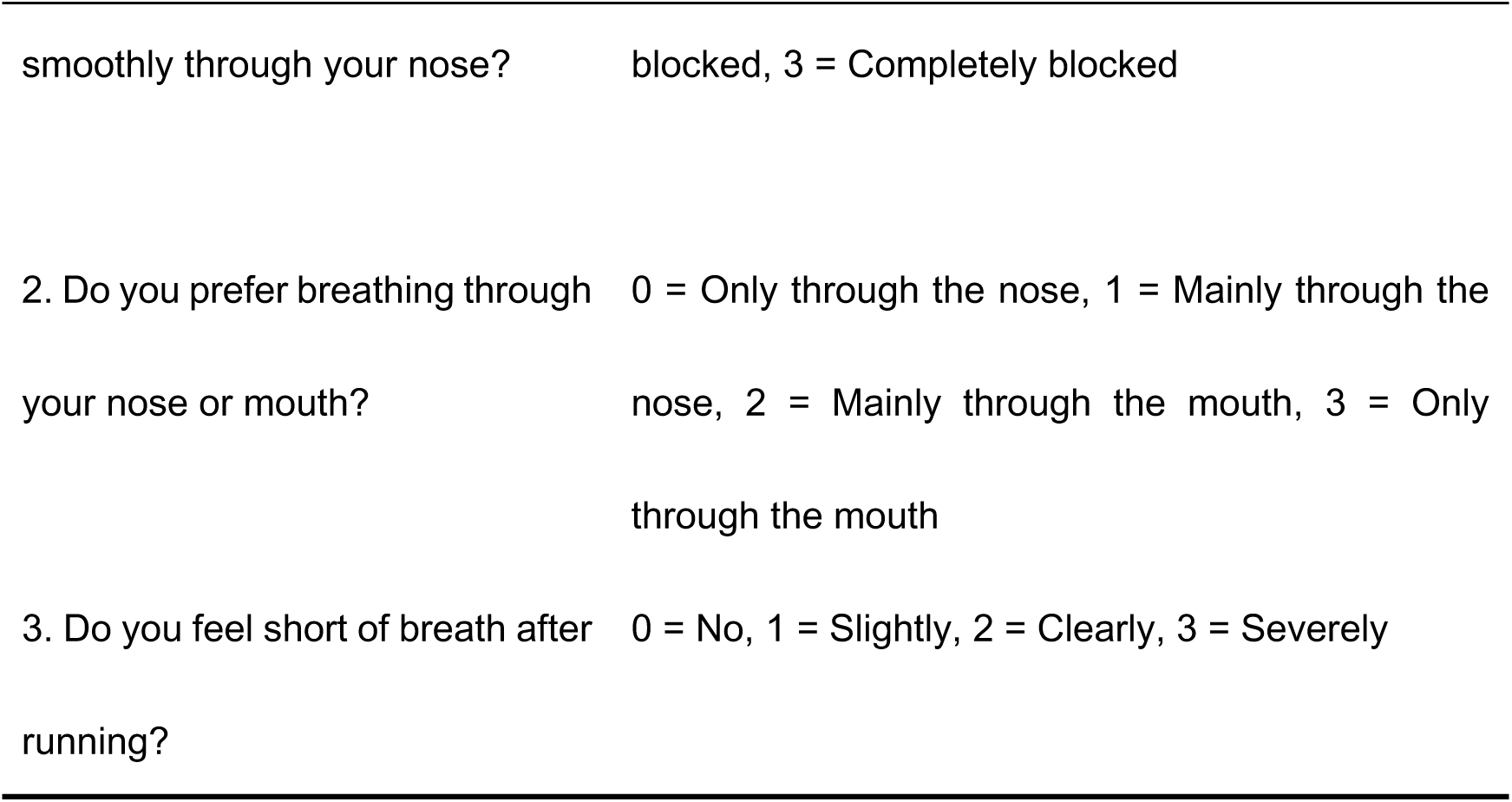
Mouth Breathing Symptom Screening Questionnaire

The criteria for the mouth breathing group were a total score of ≥6 on the questionnaire and a symptom duration of ≥6 months. The criteria for the nasal breathing group were a total score of <3 on the questionnaire and no symptoms of mouth breathing.

### CBCT imaging

Positioning: the research subjects sat upright after putting on lead aprons, with the head fixed so that the orbital-auricular plane was parallel to the ground and the mid-sagittal plane of the face was perpendicular to the ground. The upper and lower teeth were in intercuspal contact, with both eyes looking straight ahead. The tongue and perioral muscles were relaxed, with steady and even breathing, and the upper and lower lips were naturally closed, without chewing or swallowing. The CBCT scan was performed using an i-Cat (Imaging Sciences International, Hatfield, PA, USA) cone-beam machine at 85 kV, 5.5 mA, a field of view of 14×10 cm, a voxel size of 0.25 mm, and a scan time of 9 seconds. All images were taken by the same technician using the same equipment under the same exposure conditions.

### CBCT data processing and measurements

All CBCT data were imported into the Mimics 21.0 software (Materialise, Belgium) in DICOM format. The coronal, axial, and sagittal planes were reconstructed in multiplanar reconstruction mode, reconstructed with a slice thickness of 0.2 mm. A three-dimensional coordinate system was positioned and adjusted so that the line connecting the anterior and posterior nasal crests (A-B) was parallel to the horizontal plane. Additionally, the nasal septum was aligned parallel to the sagittal plane. The pharynx was divided into three segments—nasopharynx, velopharynx, and hypopharynx—based on the following landmark points. For the nasopharynx, the anterior boundary was defined by a vertical line drawn from the posterior nasal crest point B perpendicular to line A-B, separating the nasopharynx from the nasal cavity. The lower boundary was defined by the extension of line A-B, the upper boundary was the cranial base, and the posterior boundary was the posterior pharyngeal wall. A line parallel to A-B was drawn from the highest point C of the nasopharynx, and the vertical distance between this parallel line and A-B was defined as the length L1 of the nasopharyngeal cavity. Regarding the velopharynx, a vertical line parallel to A-B, drawn from the lowest point D of the uvula, served as the lower boundary. The vertical distance between this parallel line and A-B was defined as the length L2 of the velopharyngeal cavity. For the hypopharynx, a line parallel to A-B, drawn from the apex of the epiglottis E, served as the lower boundary. The vertical distance between this line and the lower boundary of the velopharynx was defined as the length L3 of the hypopharyngeal cavity (Figure 1). The total length of the pharynx (L) was calculated as L = L1 + L2 + L3.

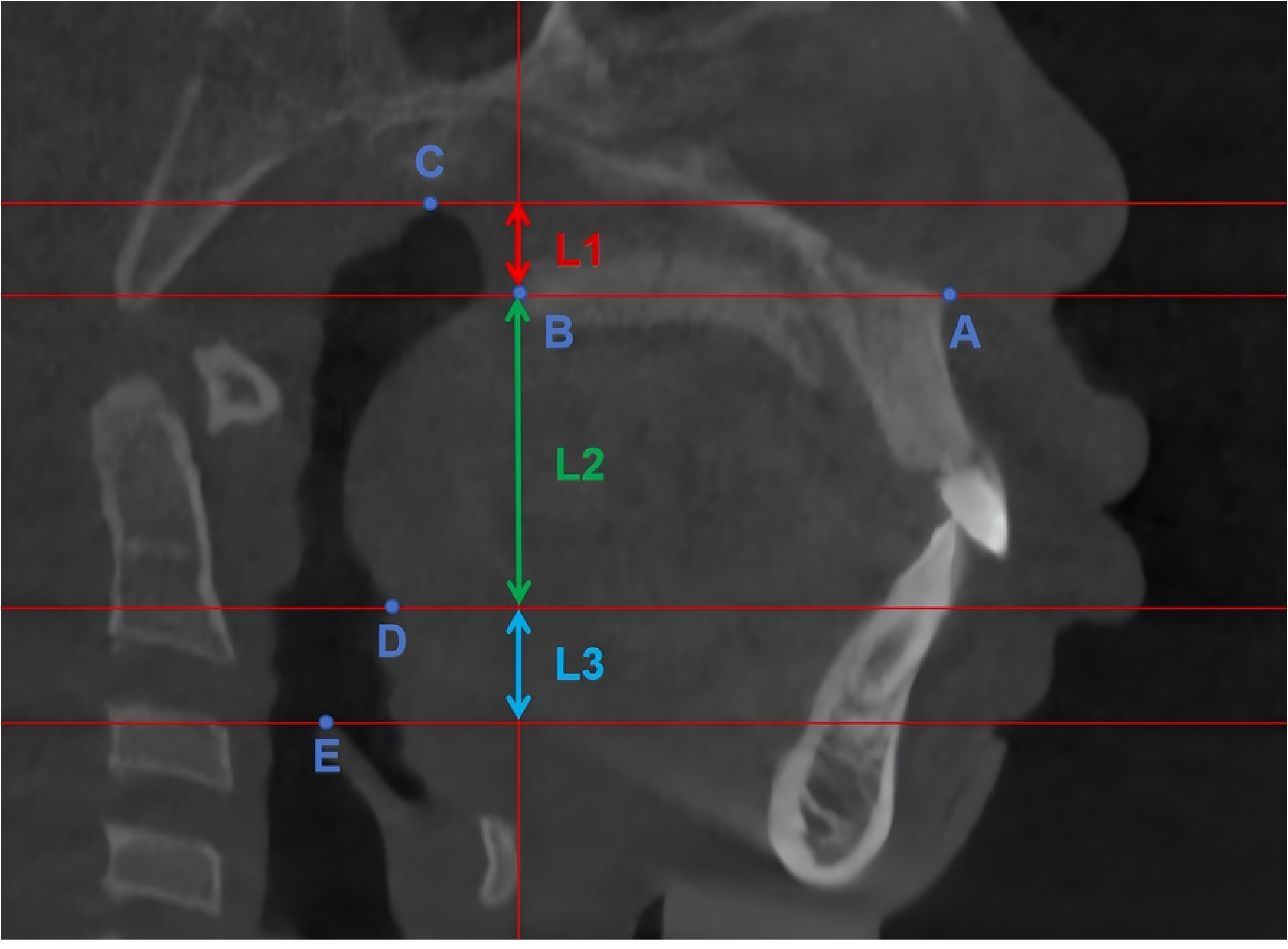

The bone tissue Hounsfield unit (HU) threshold ranged from 226 to 3071, and the soft tissue structure ranged from -1024 to 226. The volumes of the nasopharyngeal (V1), velopharyngeal (V2), and hypopharyngeal (V3) cavities were respectively extracted according to the above segmentation method (Figure 2). The total volume of the pharynx (V) was calculated as V = V1 + V2 + V3.

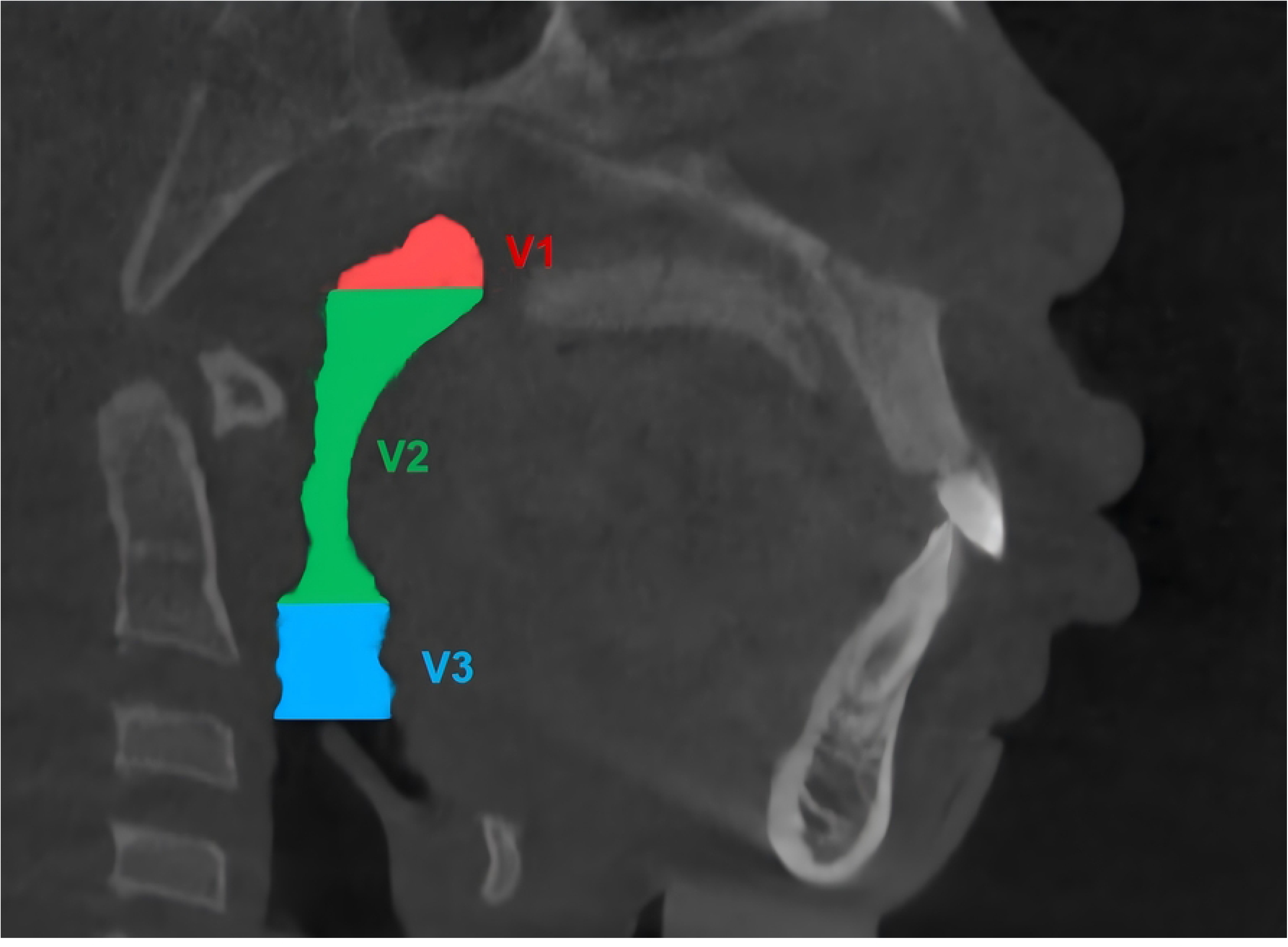

The linear density (LD) of the pharyngeal cavity volume represents the volume per unit length, calculated by the formula LD = V/L. Similarly, LD1, LD2, and LD3 represent the linear densities of the nasopharyngeal, velopharyngeal, and hypopharyngeal cavity volumes, respectively.

### Statistical analysis

All indicators in this study were measured three times by the same observer, and the average value was taken as the final value of each observation indicator for statistical analysis. The data were entered into SPSS 21.0 software. A 95% confidence level was used, and the significance level was set at 0.05. Continuous measurement data were expressed as mean ± standard deviation. Independent-samples t-tests were used for comparisons between the two groups, and chi-square tests were used for count data.

## RESULTS

Table 2 presents the comparison of general data of the study subjects. Sixty-two children were divided into two groups based on their breathing patterns: 30 in the mouth breathing group, including 22 males and 8 females, and 32 in the nasal breathing group, including 20 males and 12 females. There were no statistically significant differences in gender, age, and BMI between the two groups (P > 0.05), indicating baseline comparability of the groups. Tables 3 and 4 compare the pharyngeal cavity volumes and heights at different sites in the two groups of children. The results show that there were statistically significant differences in the total pharyngeal cavity volume and the volumes of the nasopharynx, velopharynx, and hypopharynx between the two groups (P < 0.05). The total pharyngeal cavity volume and segmental volumes in the nasal breathing group were larger than those in the mouth breathing group. However, there were no significant differences in the segmental and total lengths of the pharyngeal cavity at the same sites between the two groups (P > 0.05). Table 5 compares the volume-to-length ratios of the total pharyngeal cavity and segmental volumes in the two groups of patients. The results show that there were statistically significant differences in these ratios for the nasopharynx, velopharynx, and hypopharynx between the two groups (P < 0.05). Based on the results, it is speculated that, normalized by length, the pharyngeal cavity cross-sectional area of mouth breathing patients is narrower.

**Table 2.**
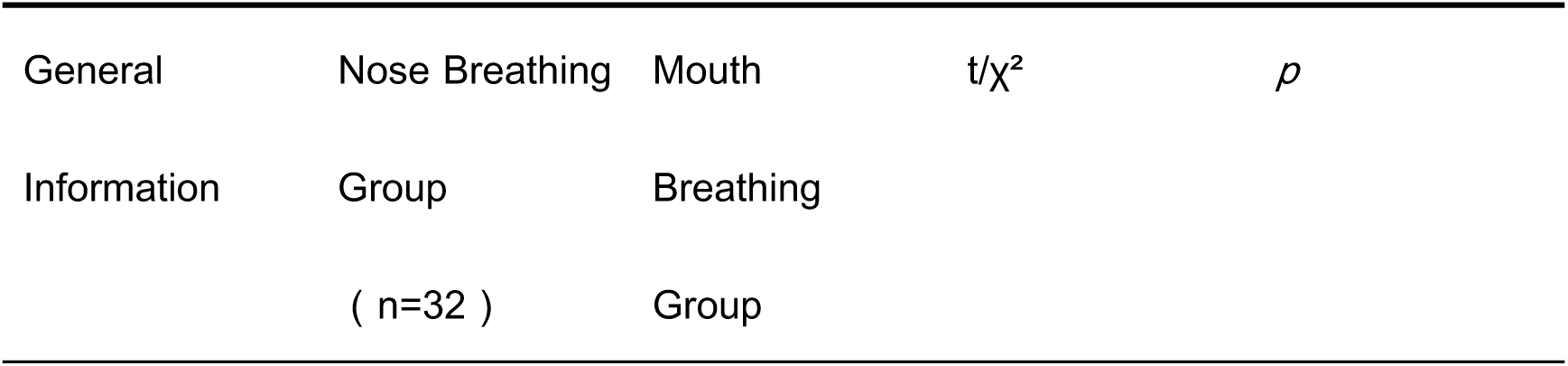
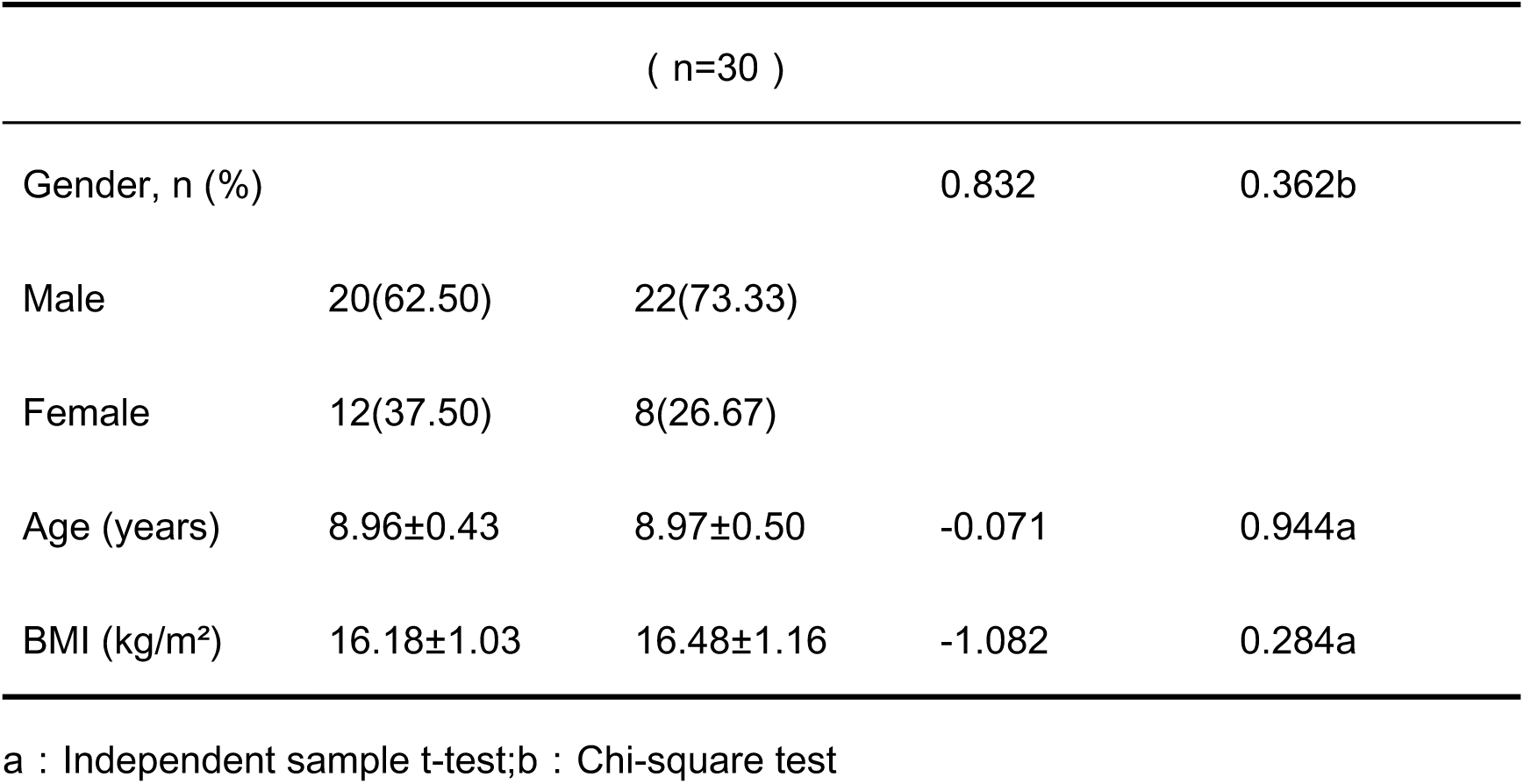
Comparison of General Information of the Research Subjects (Mean ± SD)

**Table 3:**
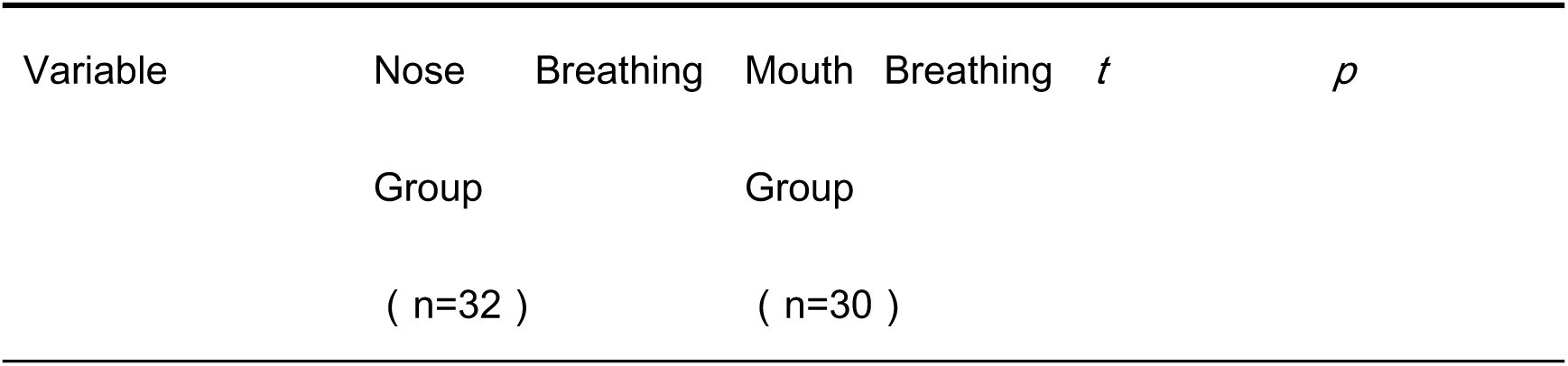
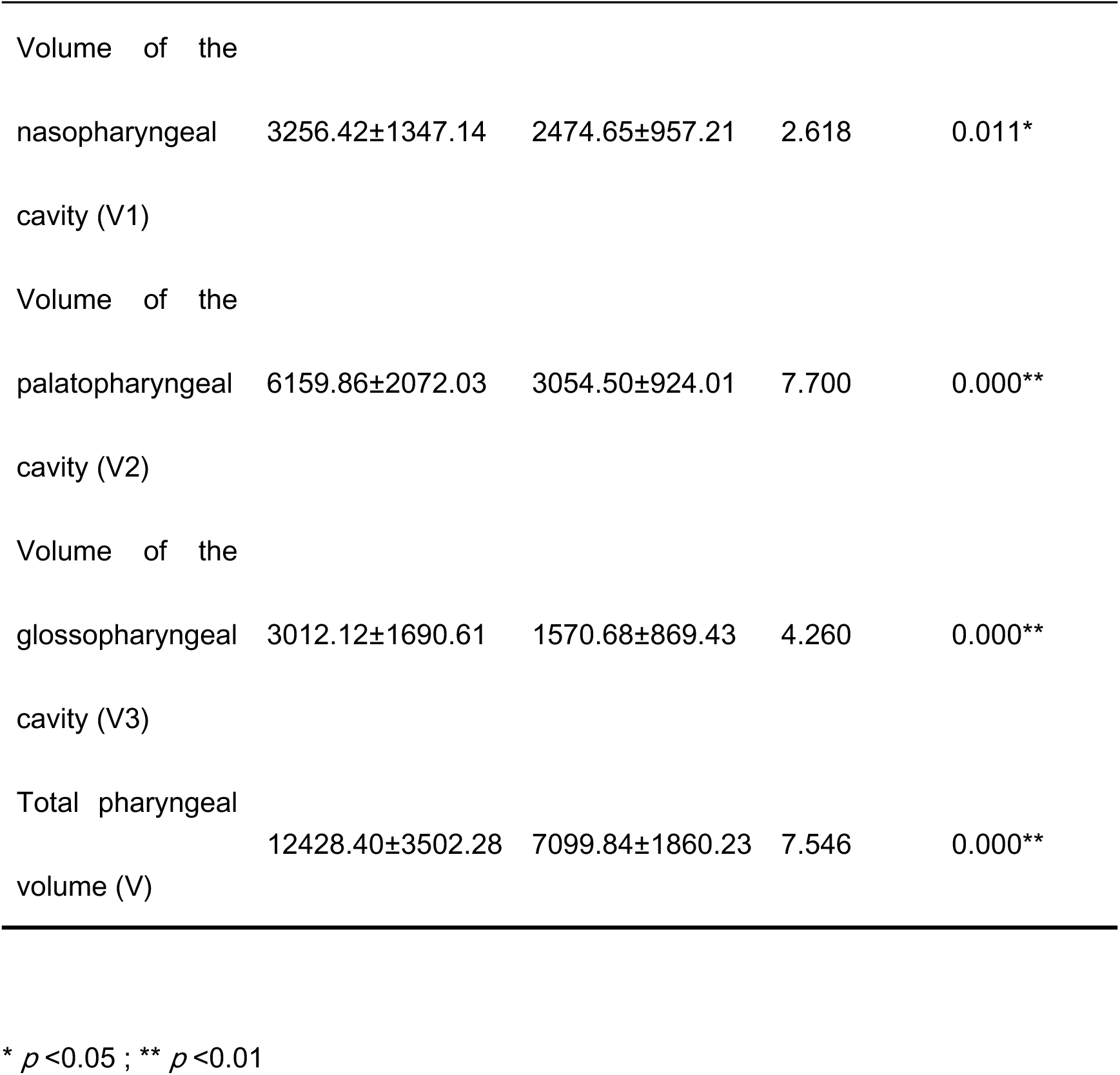
Comparison of pharyngeal cavity volumes (mean ± standard deviation)

**Table 4.**
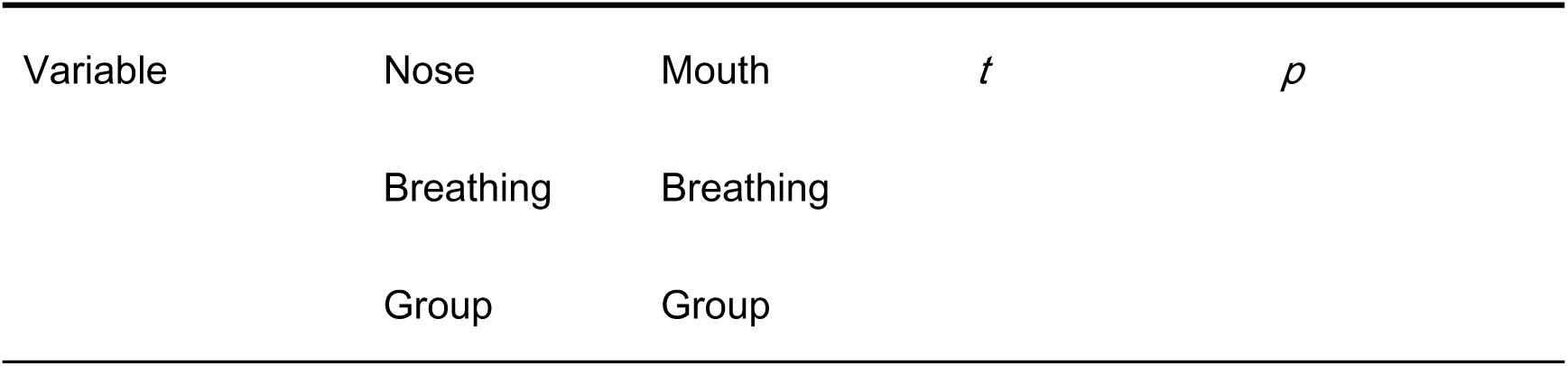
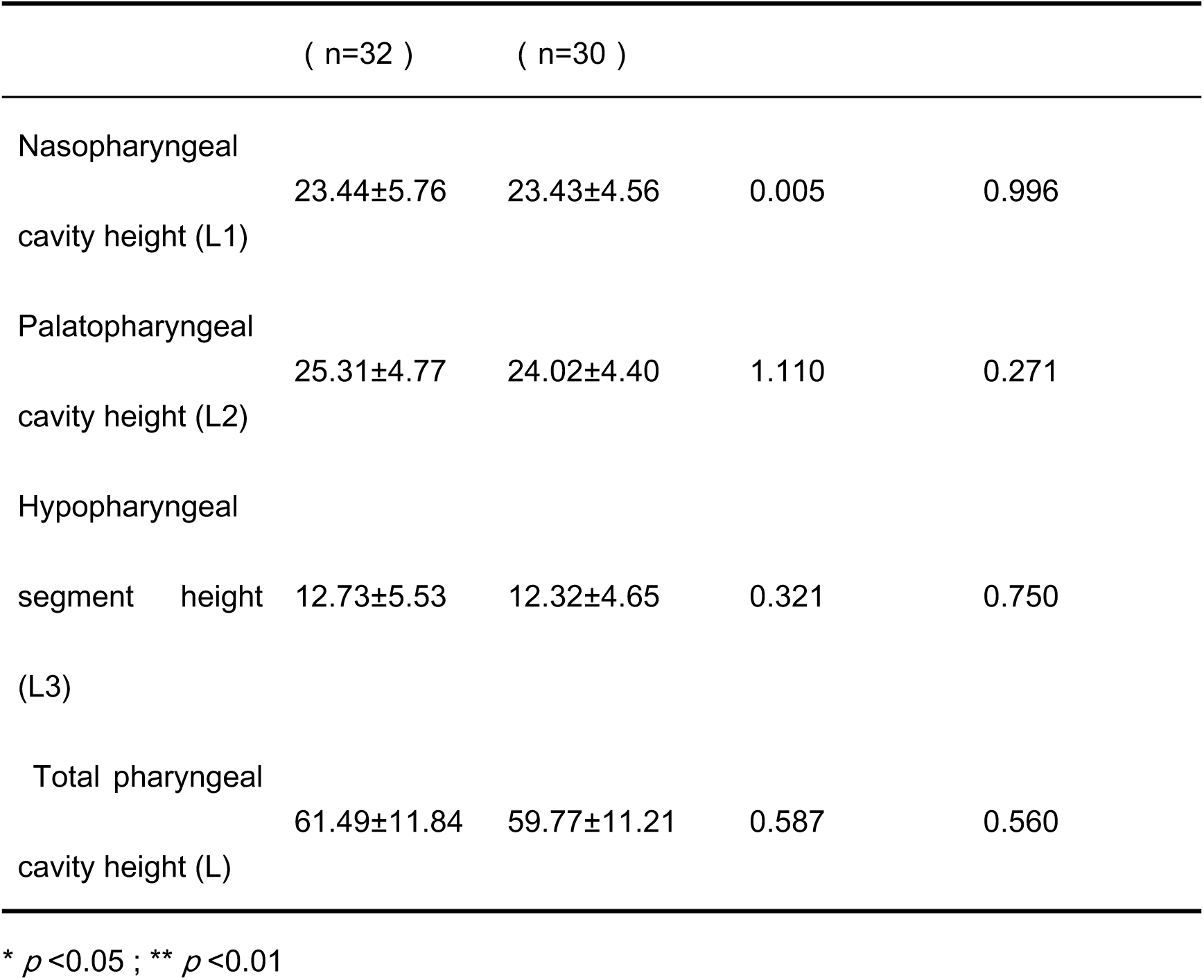
Comparison of pharyngeal cavity heights (mean ± standard deviation)

**Table 5.** Comparison of linear density of pharyngeal cavity volume (mean ± standard deviation)

| Variable | Nose | Mouth | *t* | *p* |
| --- | --- | --- | --- | --- |
|  | Breathing | Breathing |  |  |
|  | Group | Group |  |  |
|  | ( n=32 ) | ( n=30 ) |  |  |
| Linear density of<br>nasopharyngeal cavity<br>volume (LD1) | 138.41±47.<br>24 | 105.71±36.68 | 3.030 | 0.004** |
| Linear density of<br>pharyngeal cavity volume<br>of the palatopharyngeal<br>segment (LD2) | 241.26±61.<br>93 | 127.58±31.00 | 9.224 | 0.000** |
| Linear density of<br>hypopharyngeal cavity<br>volume (LD3) | 239.16±79.<br>14 | 127.32±43.05 | 6.970 | 0.000** |
| Linear density of total<br>pharyngeal volume (LD) | 201.63±40.<br>66 | 119.01±22.38 | 9.993 | 0.000** |

\* $p < 0.05$ ; \*\* $p < 0.01$

## DISCUSSION

Mouth breathing is essentially a compensatory behavior of the body in response to increased ventilation resistance ^[20, 21]^. Its causes are diverse and mainly include the following categories ^[22]^: anatomical stenosis, such as nasal obstruction, pharyngeal stenosis, and laryngeal abnormalities ^[23]^; functional causes, such as abnormal central respiratory drive, positional ventilation disorders, and upper airway collapse related to obesity ^[24]^; developmental factors, such as abnormal vertical facial bone type and compensatory posterior displacement of the tongue due to low hyoid bone position ^[25]^; and habitual factors, such as abnormal neuromuscular respiratory function caused by neuromuscular dysfunction ^[26, 27]^.

Given these causes, this study chose pharyngeal cavity morphology as the focus for the following reasons. Firstly, the pharynx is a high-incidence area of upper airway obstruction in 8- to 10-year-old children, and adenoids and tonsillar hypertrophy are highly coincident with this age group ^[28]^. Secondly, CBCT can clearly display bony landmarks and soft tissue boundaries. Additionally, Mimics 3D reconstruction can accurately quantify the morphological changes in this area. Moreover, pharyngeal stenosis may either induce mouth breathing, such as adenoid hypertrophy forcing children to breathe through the mouth, or it may be the result of mouth breathing, such as long-term mouth breathing causing the tongue to fall back, thereby exacerbating pharyngeal narrowing ^[29–31]^. Previous studies mostly used a single parameter, such as the minimum cross-sectional area of the airway or airway volume, to evaluate the degree of stenosis. However, aerodynamics indicates that ventilation resistance not only depends on the cross-sectional area at a certain point but is also closely related to the length and morphological continuity of the stenotic segment ^[23, 32, 33]^. "Pharyngeal volume linear density" is a measurement index first proposed in this study, which refers to a continuous quantitative gradient of pharyngeal volume per unit length. This index not only reflects the degree of local stenosis but also can evaluate the compensatory differences in different pharyngeal subregions.

This study revealed significant differences in pharyngeal structure between mouth-breathing and nasal-breathing children through three-dimensional morphological analysis. The results showed that the volumes of the nasopharynx, velopharynx, and hypopharynx in the mouth-breathing group were significantly smaller than the corresponding volumes in the nasal-breathing group, while there was no statistically significant difference in pharyngeal length parameters between the two groups. This phenomenon suggests that the three-dimensional morphological changes in the pharynx associated with the mouth-breathing pattern mainly manifest as narrowing in the transverse or anteroposterior direction rather than longitudinal elongation. Consequently, there is a significant decrease in pharyngeal volume relative to length. This finding provides a new anatomical perspective for understanding the impact of mouth breathing on facial development.

From a biomechanical perspective, the general reduction in pharyngeal volume may be related to compensatory soft tissue remodeling associated with the mouth-breathing pattern. Long-term mouth breathing may cause tongue posterior displacement, decreased soft palate tension, and thickening of the lateral pharyngeal soft tissue. These adaptive changes maintain basic ventilation function while compressing the three- dimensional space of the pharynx ^[34, 35]^. Furthermore, the significant narrowing of the hypopharynx volume may indicate a downward displacement of the hyoid bone and an increase in the volume of the posterior tongue ^[31, 36]^. Moreover, the reduction in nasopharyngeal volume may be directly related to the anteroposterior narrowing of the nasopharyngeal airway caused by adenoid hypertrophy, while the narrowing of the velopharynx may reflect the impact of tonsillar hypertrophy or abnormal tension of the velopharyngeal muscle group on the lateral space.

From the perspective of clinical translation value, the general narrowing of the three-dimensional pharyngeal morphology may form a vicious cycle. The narrowed pharyngeal structure increases respiratory resistance, prompting children to rely more on the mouth breathing pattern, and continuous mouth breathing further aggravates the adaptive remodeling of soft tissue, ultimately leading to abnormal development of the maxillofacial skeleton ^[2, 29, 37, 38]^. This mechanism provides a new explanation for the formation of "mouth breathing face." The restricted pharyngeal space may change the position of the tongue and the growth direction of the mandible, which induces mandibular retrusion and excessive vertical development ^[15, 36]^. The study suggests that mouth breathing intervention should be designed individually based on the morphological characteristics of different pharyngeal subregions. For instance, patients with nasopharyngeal segmental stenosis should focus on evaluating adenoid hypertrophy. In contrast, those with significantly reduced volume in the glossopharyngeal segment need to pay attention to the size of the tonsils and combine tongue positioning training or mandibular repositioning correction.

### Research strengths and limitations

This study adopted a six-month prospective follow-up to reduce the bias of misclassification. This design is superior to most cross-sectional studies. Secondly, this study first proposed the concept of "pharyngeal volume linear density" to more accurately quantify the degree of local pharyngeal stenosis, providing a new analytical paradigm. However, the sample size of this study is relatively limited, and the age is mainly concentrated between 8 and 10 years old, which may limit the general applicability of the results. Additionally, this study used three-dimensional measurement technology to improve the accuracy of pharyngeal space assessment. However, the dynamic impact of breathing patterns on pharyngeal morphology, especially the volume changes caused by different breathing phases (inhalation and exhalation), still needs to be further explored in combination with functional tests.

### Summary and outlook

This study provided objective imaging markers for the early detection of mouth breathing through quantitative analysis of the three-dimensional morphology of the pharynx. Although the etiology may involve interactions among multiple bodily systems, the study focused on the core sequence of local pharyngeal stenosis, compensatory mouth breathing, and atypical maxillofacial development. This focus offers a practical entry point for clinical intervention. The expected results not only establish objective quantitative standards for the early detection of high-risk children in clinical practice but also develop a measurement framework that serves as a methodological foundation for subsequent large-scale data research. Future studies should clarify the dynamic patterns of pharyngeal morphological changes and their long-term effects on overall health through longitudinal cohort tracking. Moreover, these structural parameters can be integrated into machine learning models to create automated screening tools based on medical imaging. Ultimately, this approach will enable a shift from empirical diagnosis to precise prediction, thereby advancing the intelligent era of children’s respiratory health management.

## CONCLUSIONS

Taken together, mouth-breathing children exhibit significantly smaller pharyngeal volumes without differences in pharyngeal length, resulting in a reduced volume-to-length ratio (cross-sectional area per unit length). This confirms a narrower three-dimensional pharyngeal morphology in mouth breathers .

## Data Availability

The data underlying the results presented in the study are available from The Second Nanning People's Hospital

